# Household Vulnerability and Coping Amid the COVID-19 Pandemic in Bamako, Mali: Assessing Sociodemographic Factors Associated with Household Hardships

**DOI:** 10.1101/2024.11.27.24318121

**Authors:** Jonathan A. Muir, Uduma U. Onwuchekwa, Zachary J. Madewell, Moussa O. Traore, Moussa Kourouma, Fatima Keiri, Solveig A. Cunningham, Karen L Kotloff, Milagritos D. Tapia, Samba O Sow

## Abstract

**Background:** COVID-19 resulted in vast disruption to life in the 21^st^ century as governments implemented containment measures to quell disease spread. We assessed knowledge of local interventions and household coping strategies used to attenuate the impact of household hardships. We further examined associations between household and community characteristics with household hardships.

**Methods:** We conducted a cross-sectional household survey between August and September 2022 through a retrospective questionnaire fielded in the Health and Demographic Surveillance System (HDSS) operating in Bamako, Mali. Logistic regression was used to analyze associations between household characteristics, government interventions, and household hardships during the pandemic.

**Results:** The most commonly reported hardships were increases in food prices and food insecurity; roughly 18% of households reported experiencing at least 1 hardship. Common coping strategies included asking for help from family or friends (55.6%). Only 2.8% of households reported seeking government assistance. Households headed by younger individuals, males, and unemployed individuals were at greater risk of experiencing hardships. Government closure of businesses was strongly associated with household hardships.

**Conclusions:** Households in the Bamako HDSS experienced a variety of hardships during the pandemic— the most prevalent hardships were increases in local food prices and food insecurity. The association between government closure of businesses and household hardships points to the need for balancing public health measures with socioeconomic considerations. Households headed by individuals with lower education and/or unemployed were at greater risk of experiencing a hardship. Future policies and interventions should target aid to households reflecting these characteristics.

## Introduction

Repercussions from the COVID-19 pandemic extended far beyond direct health consequences.(1) Governmental measures like lockdowns and curfews, implemented to control the spread of SARS-CoV-2 (the virus that causes COVID-19), triggered a cascade of social and economic disruptions.(2, 3) These interventions, while helpful for curbing SARS-CoV-2 transmission, also led to economic strain, restricted access to food and healthcare, and pushed vulnerable populations towards food insecurity and health risks.(4, 5, 6, 7, 8) Businesses, including healthcare providers, were forced to close or limit services, potentially jeopardizing access to essential care and resources (including sources of income) for vulnerable populations in the community. Reduced access could have inadvertently amplified the virus’s direct effects by increasing vulnerabilities to undernutrition and other risk factors for severe illness. However, limited population-based surveillance in resource-scarce settings has hampered our understanding of the extent of resource restrictions and hardships experienced during the pandemic. This is particularly the case for Mali, one of the poorest countries in the world with some of the highest levels of child mortality and lowest life expectancy, where there remains a paucity of data on socioeconomic impacts of the pandemic.

Vulnerability, broadly defined, is the cumulative effect of cultural, economic, institutional, political, and social processes that amplify socioeconomic differences in the experience of and recovery from hazards.(9, 10) In the context of disasters, it is often not the hazard itself that creates the crisis; rather, the crisis is the impact on individual and community coping patterns and on the inputs and outputs of social systems.(11, 12, 13, 14) Social vulnerability is the result of social disparities that shape or influence the susceptibility of different groups to hazards, while also limiting their capacity to respond.(9, 15) Factors often associated with vulnerability include demographic characteristics such as age, ethnicity, race, and sex; socioeconomic status (e.g., income, wealth, employment, education); household composition (e.g., presence of children or elderly); and built environment (housing and transportation).(16) Social vulnerability also involves place disparities stemming from community characteristics. For example, differential availability of scarce resources between urban and rural areas may exacerbate vulnerabilities to hazards.(15, 17, 18, 19, 20) To understand the broader consequences of the pandemic, it is important to consider economic, political, and social markers of vulnerability at both the individual and community level.(21)

The arrival of COVID-19 in Mali, like in many other parts of the world, unleashed a wave of disruptions exceeding the immediate health threat. On March 18^th^, 2020, prior to its first confirmed COVID-19 case, Mali enacted preventative measures like flight suspensions, school closures, and public gathering bans.(22, 23) Following the first cases on March 25^th^, a state of emergency was declared with a curfew implemented.(24) By March 28^th^, with the first death recorded, Mali had restrictions comparable to North America and Western Europe.(22, 23) Mali’s pre-existing vulnerabilities, including poverty, food insecurity, limited healthcare access, a fragile agricultural sector, climate sensitivity, dependence on regional ports, and ongoing conflict in some regions, likely exacerbated the pandemic’s impact.(25, 26) These vulnerabilities were further exacerbated by restrictions on movement and overwhelmed healthcare systems, leading to increased food insecurity and jeopardizing the health of vulnerable groups like children, pregnant women, and mothers. Data limitations hinder a comprehensive understanding of these hardships and their differential effects. Investigating the long-term consequences of these interventions within the context of Mali’s unique challenges can inform future pandemic preparedness and response strategies tailored to such settings.

Furthermore, understanding the level of knowledge about COVID-19 prevention measures among Malian households is important for assessing the effectiveness of public health messaging and interventions. Previous studies in sub-Saharan Africa have demonstrated positive associations between knowledge of COVID-19 symptoms, SARS-CoV-2 transmission, and prevention, and the implementation of public health measures like hand hygiene, social distancing, and facemask use.(27, 28) Additionally, knowledge of these measures has been linked to higher vaccination coverage, one of the most effective strategies for protecting individuals against COVID-19 hospitalization and death.(29, 30, 31, 32) Understanding knowledge levels is useful for designing effective public health messaging and interventions tailored to the population context and needs.

This study leverages data from an existing Health and Demographic Surveillance System (HDSS) in Mali to analyze the prevalence of hardships experienced during the pandemic as well as knowledge about COVID-19 prevention measures held by Malian households. We focus on key areas such as economic insecurity, food insecurity, healthcare access, and education disruptions, along with exploring household responses to these hardships and their association with knowledge levels.(25, 26)

## Methods

### Study Design

This cross-sectional study is part of a larger examination within the Child Health and Mortality Prevention Surveillance (CHAMPS) network to assess implications and potential consequences of pandemic-related measures, including lockdowns, social distancing, and related policies aimed at stemming the transmission of SARS-CoV-2.(6, 7, 8, 33, 34) Specifically, the data were gathered to increase understanding of how these measures may have contributed to broader household socioeconomic challenges. This includes potential impacts on access to healthcare, food availability, and livelihoods, among households, with particular focus on vulnerable groups such as those with children and pregnant women residing in the Bamako HDSS.(35)

### Study Setting

Mali is a large, predominantly arid country in West Africa, covering over 1,241,238 square kilometers (∼479,000 sq. mi) with a population of approximately 21.4 million. The population is predominantly Muslim, with a young age structure (nearly 50% under 15 years), high adult illiteracy (64.5%), high adolescent fertility, and limited access to healthcare.(36) Mali’s development and health indicators rank among the lowest in the world according to the United Nations index.

In 2002, the Centre pour le Développement des Vaccins du Mali (CVD-Mali) was established as a partnership between the Mali Ministry of Health and the University of Maryland, Baltimore, USA. CVD-Mali is a legal government entity working under the Ministry of Health. Since its inception, CVD-Mali has been dedicated to describing the epidemiology of infectious diseases, supporting the development of vaccines, and training a team of local researchers. In 2006, CVD-Mali established an HDSS in two quartiers (neighborhoods) in Bamako. One neighborhood, Banconi, is situated in Commune 1 and is in the eastern part of the city, north of the Niger River, and occupies an area of 6.23 km^2^. In 2022, the population was estimated at 153,490. The second neighborhood is Djikoroni-para; it has a surface area of roughly 5 km^2^. It is situated in Commune 4 and is in the western part of Bamako. The estimated population was 82,422 in 2022. Both neighborhoods are densely populated, and the population largely subsists on small business activity, government employment, foreign remittances, and trade. The Bamako HDSS generally conducts biannual demographic data collection rounds as part of its surveillance.

### Participants and Recruitment

For this auxiliary study, participants were recruited between August and September 2022. Study participants comprised a qualifying representative (the household head or an adult household member with good knowledge about the household). All households within the Bamako HDSS were eligible for selection into the study, which was implemented during regular HDSS round visits. We used proportion to population stratified random sampling to identify a random sample of 454 households stratified across the two communities covered by the HDSS. Simple random sampling was executed within the two catchment areas (strata) with the goal of obtaining samples from the two strata that were proportionate to the population of households residing therein—as all households actively participate in broader HDSS activities, we obtained a 100 percent response rate. Ultimately, we collected data from 317 households residing in Banconi and 137 households residing in Djikoroni, representing 69.8% and 30.1% of the sample respectively. The study adopted the definition of households and household members used by the HDSS.

### Survey Instrument

The survey instrument was designed following established methods,(37) which included identifying key concepts, framing research questions, conducting literature reviews, and drawing from preexisting questionnaires, such as the “High-Frequency Mobile Phone Surveys of Households to Assess the Impacts of COVID-19” standardized survey instrument prepared by the World Bank for assessing direct and indirect socioeconomic consequences of the COVID-19 pandemic.(38) A standardized survey instrument was prepared in English for implementation across the CHAMPS network, with adaptations for local contexts. The final questionnaire was organized into six sections: respondent identification, SARS-CoV-2 knowledge and practices; histories of symptoms; contacts with people that have COVID-19 symptoms; household economic impacts of COVID-19; and the impacts on healthcare access for household members (see supplementary materials). The instrument did not seek to ascertain COVID-19 cases. The survey prompted participants to reflect on household experiences since March 2020.

### Data Collection and Quality Assurance

Data collection was conducted electronically using tablets programmed with RedCAP,(39) with both English and French translations of the instrument presented at the same time. Communication with study participants was in their preferred language. If a fieldworker could not communicate in the preferred language of the respondent, an interpreter was sought from within the household, or the household was revisited at another time by a fieldworker who could communicate in the preferred language. Data cleaning adhered to standard procedures for the HDSS;(40, 41) data collectors and supervisors were trained on the study objectives, data confidentiality, and data collection techniques. Data quality concerns such as data inconsistencies, implausible values, or incomplete data were identified and flagged for reconciliation, which was completed by data collectors through return visits to the household.

### Statistical Analysis

#### Measures

The primary outcome variable of interest was a dichotomous indicator for whether a household reported experiencing at least one type of hardship since the onset of the COVID-19 pandemic (i.e., during the 1 to 1 1⁄2 years since March 2020). Information on these household hardships were gathered from the following survey question: “Has your household been affected by any of these events since mid-March (2020)?” Responses included job loss, business closure, increases in the price of the main food products consumed, increases in the price of agricultural or commercial inputs, and having a household member detained (see Figure 1).

**Figure 1.**
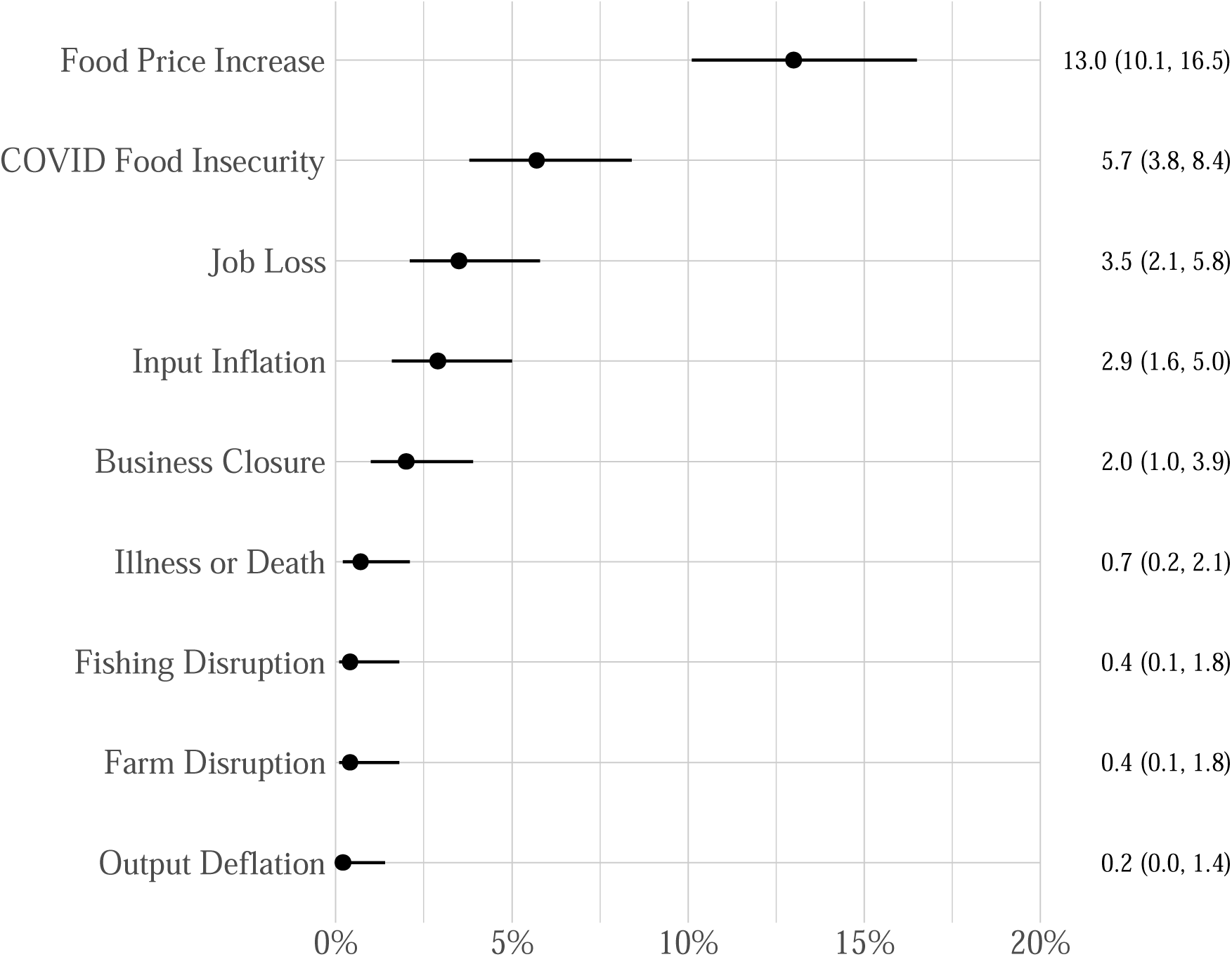
Percentage of Households that Reported a Given Hardship (n = 454).

Additional descriptive analyses were performed to describe government responses to COVID-19 and household coping strategies to experiencing any of the hardships listed above. These variables were based on data collected through the following questions: “What steps has your community/government taken to curb the spread of the coronavirus in your area?” and “How did your household cope with these difficulties [hardships] encountered since mid-March (2020)?” Corresponding answers related to government responses included advised citizens to stay home, advised citizens to avoid gatherings, closure of non-essential businesses, closure of schools and universities, curfew/lockdown, disinfection of public places, established isolation centers, sensitization/public awareness, travel restrictions, and other. Responses related to household coping strategies included borrowed from family or friends, delayed payments, generated additional income, government help, help from family or friends, NGO help, reduced food consumption, reduced non-food consumption, relied on savings, requested a pay advance, sold assets, sold harvest, took out a loan, and other. For each question, data collectors were instructed to select all applicable response categories; each response category was recoded into a unique variable coded as 0 = No (reference), 1 = Yes.

Right-hand side variables. Head of household variables were age (coded as 1 = <40 (reference), 2 = 40 to 49, 3 = 50 to 59, 4 = 60+, 5 = Don’t Know); sex (coded as Female = 0 (reference) and Male = 1); education (coded as 1 = None (reference), 2 = Primary, 3 = Secondary/Post-Secondary, 4 = Don’t Know); occupation (coded as 1 = Unemployed (reference), 2 = Professional, 3 = Sales, 4 = Manual Labor); ethnicity (coded as 1 = Bambara (reference), 2 = Fula/Malinke/Senufo, 3 = Songhai/Songinke/Tuareg, 4 = Other); and marital status (0 = Not Married (reference), 1 = Married; original marital status categories combined in the reference category included divorced, single/never married, widowed, separated, and other). Household variables included household size (coded as 1 = 1-4 (reference), 2 = 5-7, 3 = 8-9, 4 = 10+) and residence (coded as 0 = Banconi (reference), 1 = Djicoroni). In preliminary analyses, we also assessed presence of either children under the age of 5 as well as adults older than the age of 65+; however, these variables were not statistically significant in any of the models that they were included in and the models performed better with their exclusion. Government responses to COVID-19 were included as dichotomous variables coded as (0 = No, 1 = Yes) depending on whether a household reported that a given response intervention was implemented in their local neighborhood. Household responses were collected using the following question: “What steps has your community/government taken to curb the spread of the coronavirus in your area?” Interviewers were instructed to read aloud the following list of possible interventions and to select all that applied: None; Advised citizens to stay at home; Advised to avoid gatherings; Restricted travel within country/area; Restricted international travel; Closure of schools and universities; Curfew/lockdown; Closure of non-essential businesses; Sensitization/public awareness; Established isolation centers; Disinfection of public places; Other; Don’t know; and Refused to respond. From these responses, the following variables were generated: stay home; avoid gatherings; travel restrictions (composite of domestic and international travel restrictions); school closures; lockdowns; business closures; public awareness; isolation centers; and disinfection.

#### Analytic Strategy

Data preparation and analysis was completed using R version 4.2.0.(42) Missing values were less than one percent of the total observations and addressed through listwise deletion in analytic models. Descriptive statistics were reported for head of household and household characteristics. Cross-tabulations combined with **χ**^2^ tests of right-hand sided variables with the dependent variable were generated to describe potential relationships between these categorical variables. Unadjusted logistic regression models were executed to further examine associations between characteristics of the head of household, household, and government responses to COVID-19 with the dependent variable household hardships. A final model adjusted association between the dependent variable and right-hand sided variables by controlling for other variables included in the model. Right-hand sided variables were retained in the final adjusted model if their inclusion had theoretical justification based on extant literature; we assessed model fit with a comparison of Akaike Information Criterion (AIC) and Bayesian Information Criterion (BIC) scores—the smallest scores were for the fully adjusted model, indicating that this model had superior fit compared to simpler models. Results from the final logistic regression model are reported as adjusted odds ratios (AOR) with 95% confidence intervals (95% CI) and visualized with forest plots.(43, 44) We explored potential effect modification of association based on location of household residence (Banconi vs. Djicoroni) through modeling interaction effects; these explorations failed to yield statistically significant results and are not reported.

## RESULTS

The median and modal age category of household heads was 50-59 years; most households (86.3%) were headed by a male household member and most household heads (87.6%) were married (Table 1). Almost half of household heads had no education and another 28.2% had a primary level education. The most common occupation categories were manual labor (34.7%) and sales (24.3%); a little over 1 in 5 household heads were unemployed. Bambara was the most common ethnic group (35.2%); Fula/Malinke/Senufo and Songhai/Songinke/Tuareg represented about 33.0% and 16.7% of households respectively. Median household size was 8-9 household members. Roughly 70% of households resided in Banconi with the remaining 30% residing in Djicoroni.

**Table 1:**
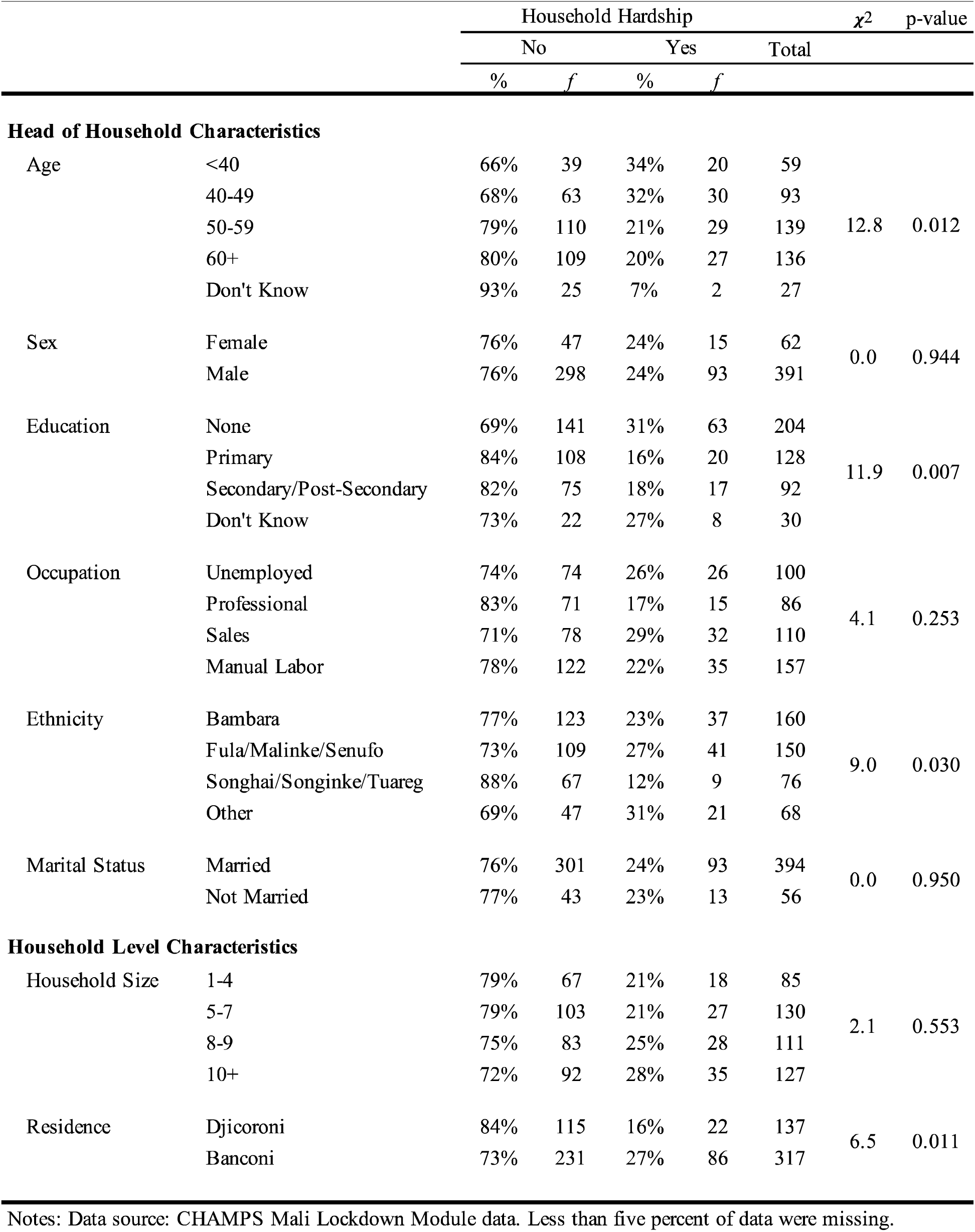
Descriptive Statistics for Head of Household and Household Characteristics (n=454)

The most common hardship reported by households was increases in food prices and food insecurity (Figure 1). However, a little less than 80% of households reported that they did not experience any of the hardships measured during the observation period; roughly 18% reported experiencing at least 1 hardship (Figure 2). The overall pattern of the number of hardships experienced during the pandemic is presented in Figure 3. Among those households that reported experiencing at least one hardship during the pandemic, the coping strategies most frequently reported were to ask for help from family or friends (55.6%) and to borrow from family or friends (47.2%). Only 2.8% of households reported seeking out government assistance and no households reported seeking out assistance from an NGO (Figure 4). Most households reported utilizing only one or two coping strategies; the maximum number of strategies implemented was four (Figure 5). With regards to government responses to COVID-19, a majority of households reported local implementation of advisories to avoid gatherings (89.9%), advisories to stay at home (85.2%), travel restrictions (64.8%), closures of schools and universities (58.4%), and curfew/lockdowns (56.6%). Less than 20% of households reported business closures (see Figure 6). A visualization of the number of government responses is presented in Figure 7 (see Table 1.1 for additional details).

**Figure 2.**
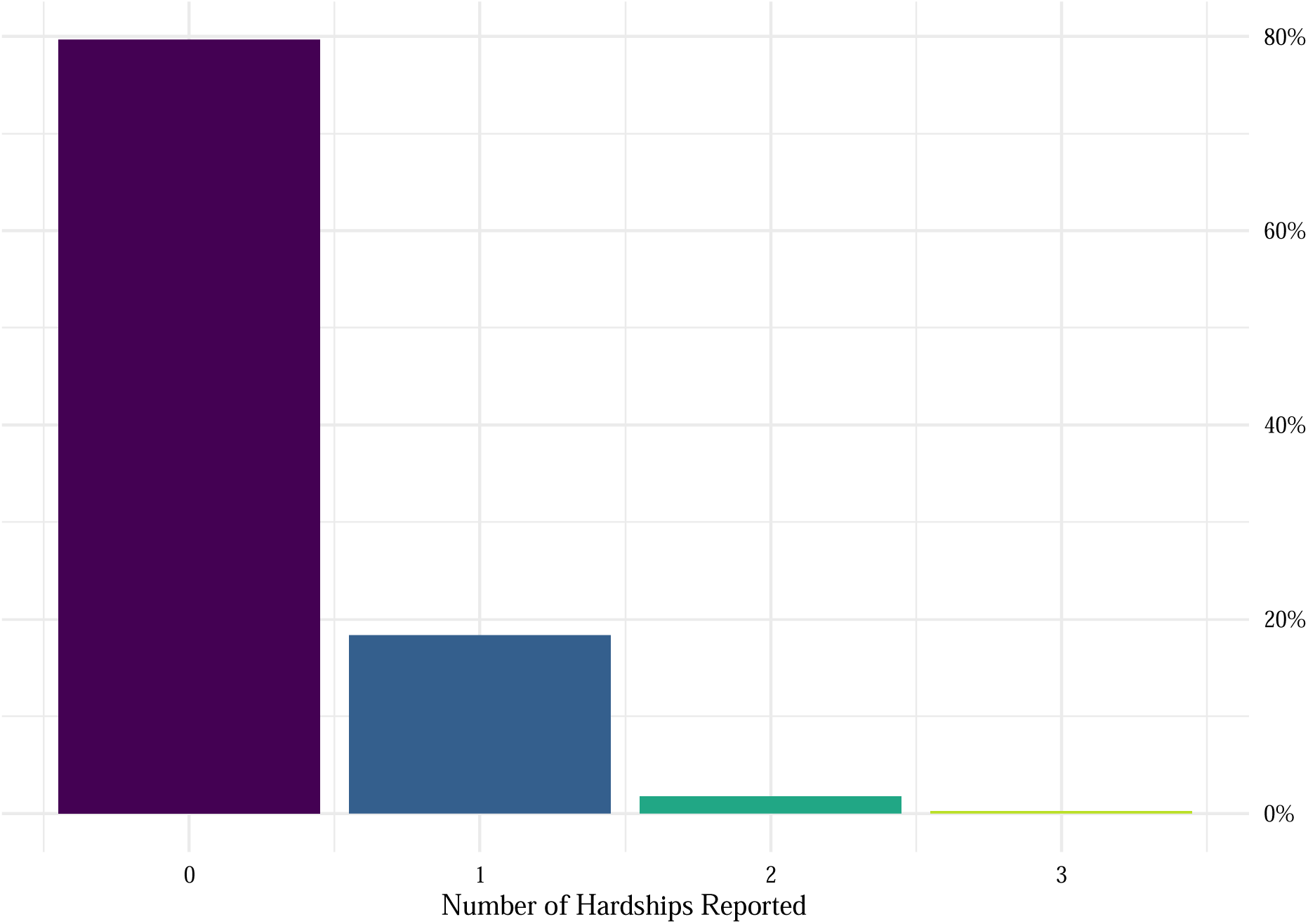
Percentage of Households that Reported a Given Number of Hardships (n = 454). Counts reflect a summation of all hardships reported by a given household; i.e., food price increase, food insecurity, job loss, input inflation, business closure, illness or death, fishing disruption, farming disruption, and output deflation.

**Figure 3.**
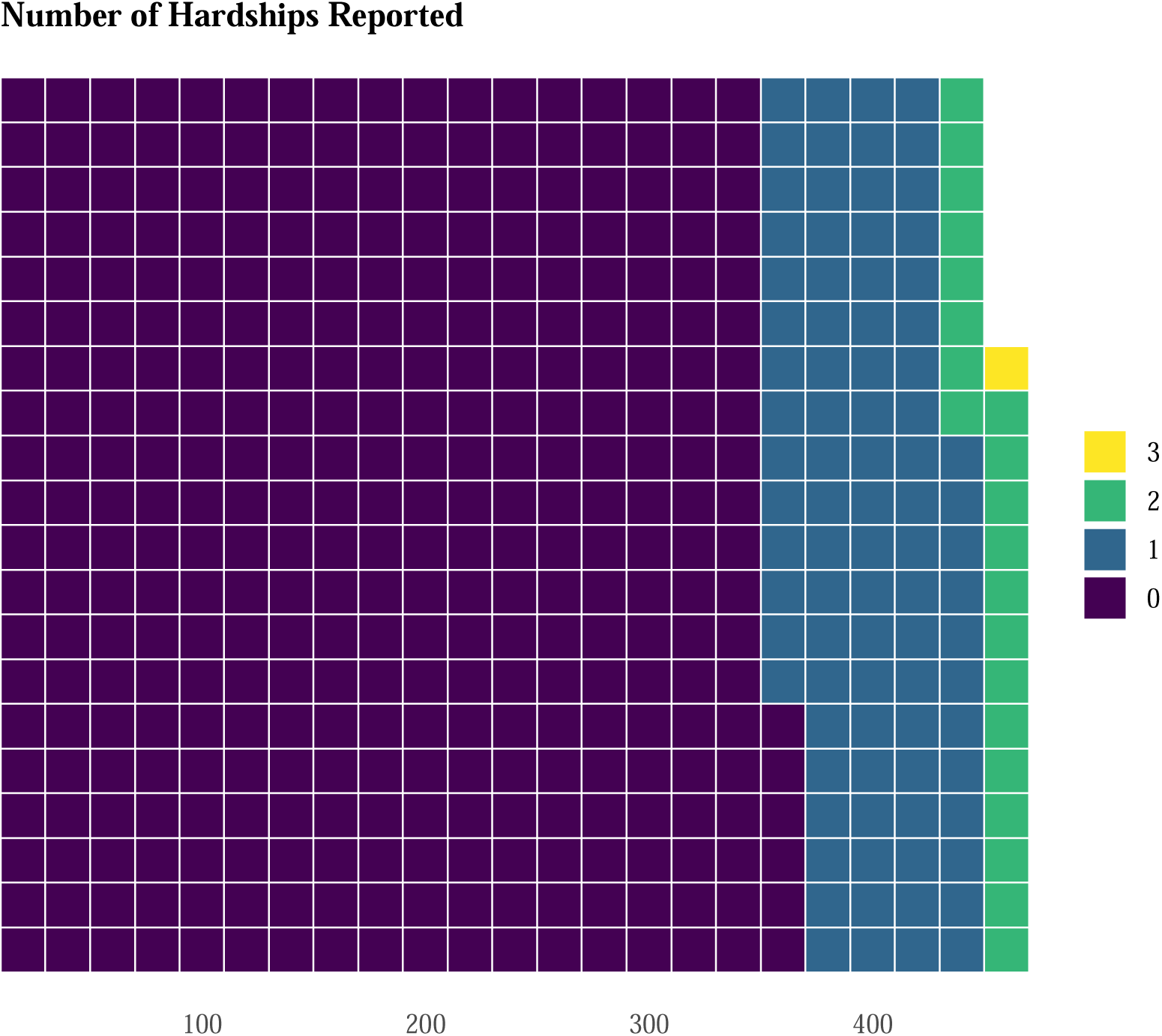
Pattern of Multiple Hardships Reported by Households (n = 454). Counts reflect a summation of all hardships reported by a given household; i.e., food price increase, food insecurity, job loss, input inflation, business closure, illness or death, fishing disruption, farming disruption, and output deflation.

**Figure 4.**
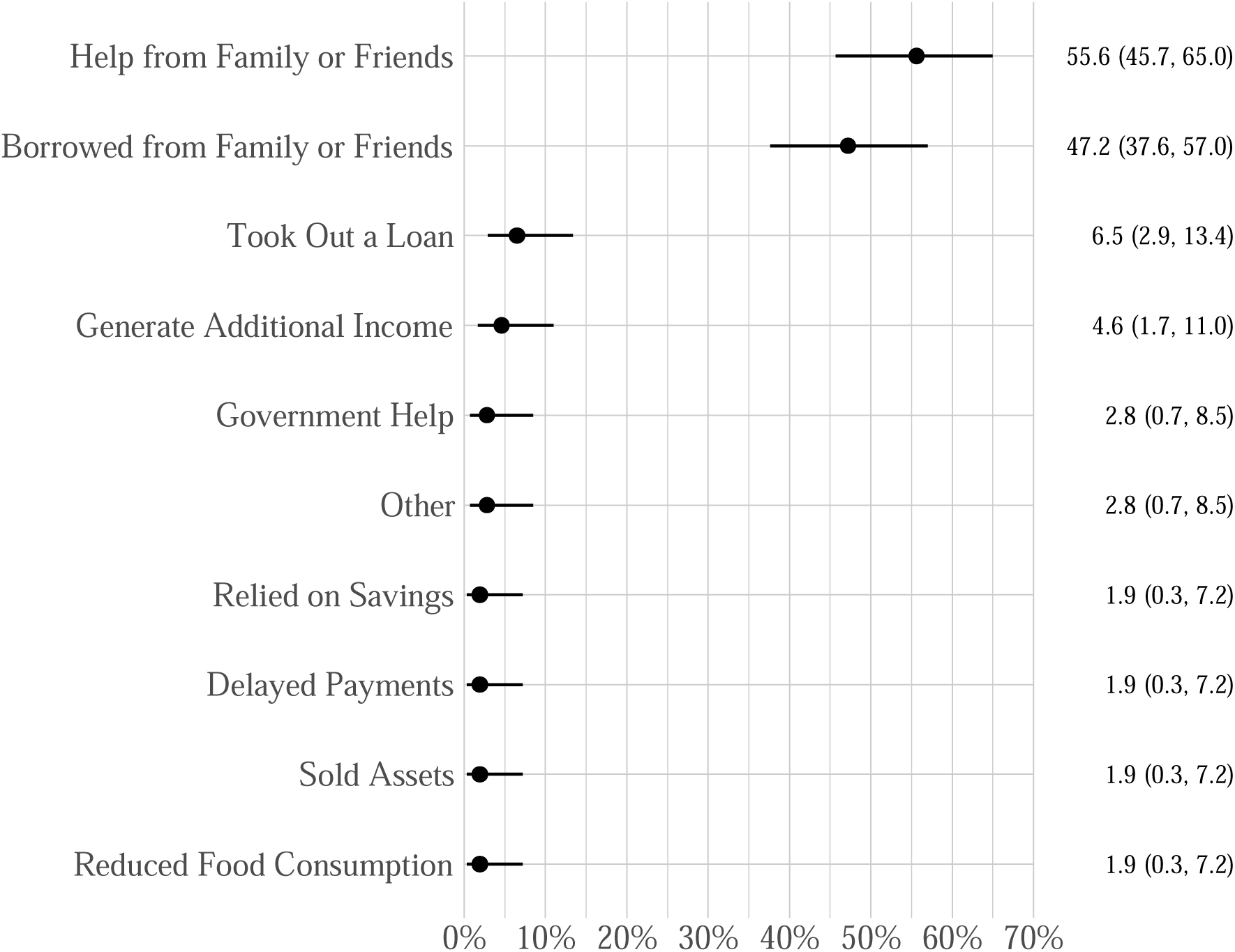
Percentage of Households that Reported a Given Response to Experiencing Household Hardships (n = 454).

**Figure 5.**
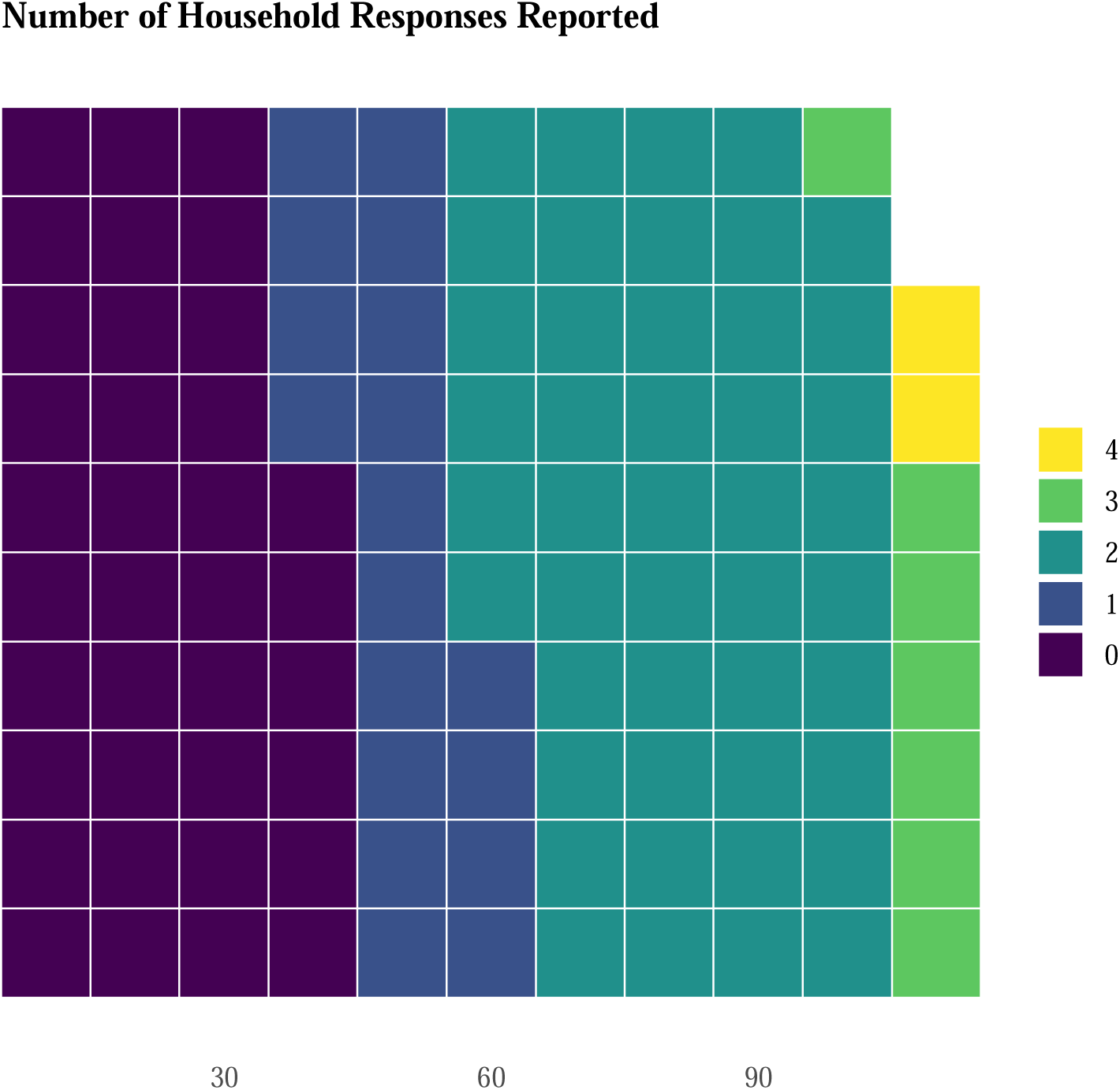
Pattern of Household Responses to Hardships (n = 454). Counts reflect a summation of all responses reported by a given household; i.e., seeking help from family or friends, borrowing from family or friends, taking out a loan, generating additional income, seeking government help, relying on savings, delaying payments, selling assets, reducing food consumption, and other.

**Figure 6.**
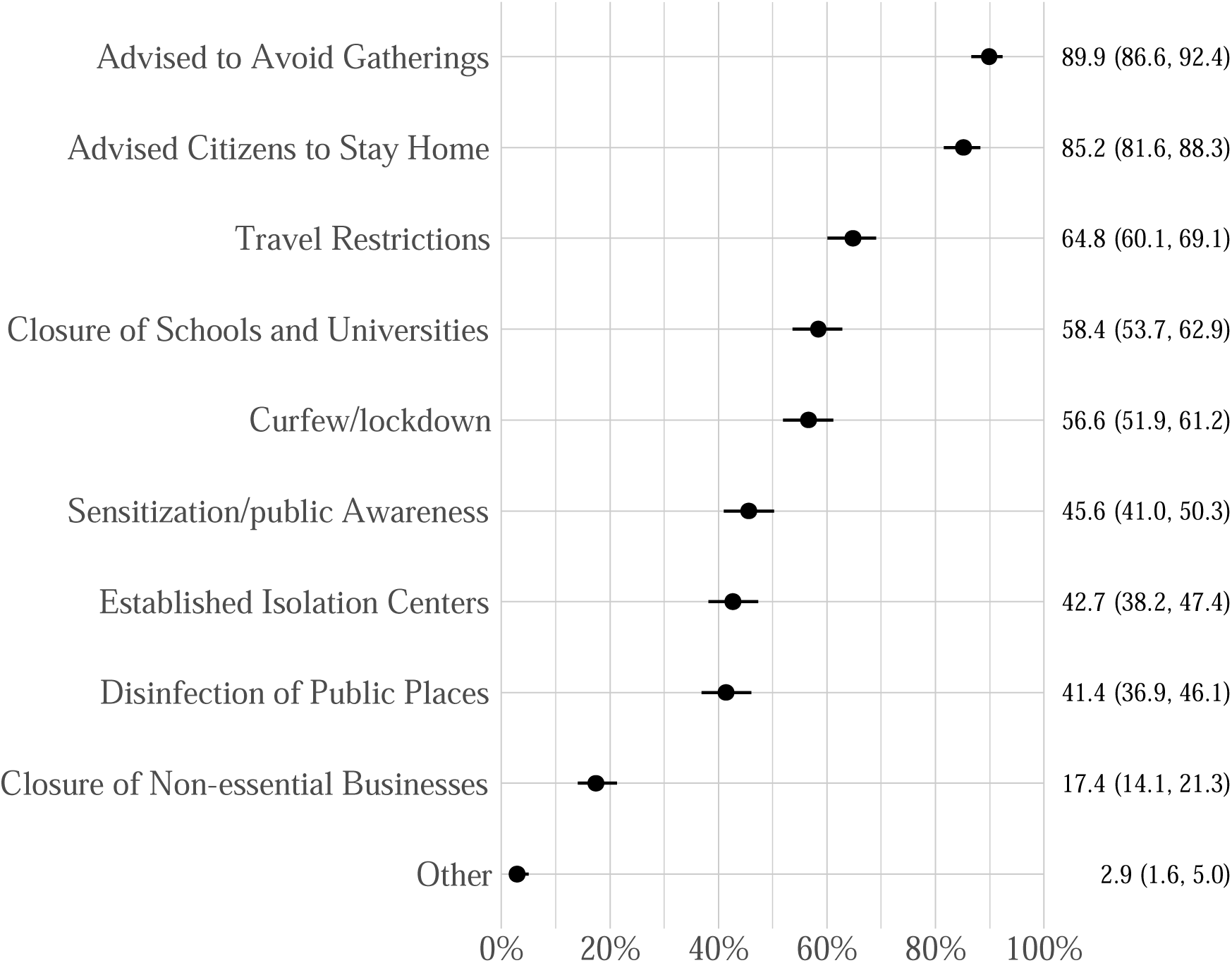
Percentage of Households that Reported a Given Government Response to COVID-19 (n = 454).

**Figure 7.**
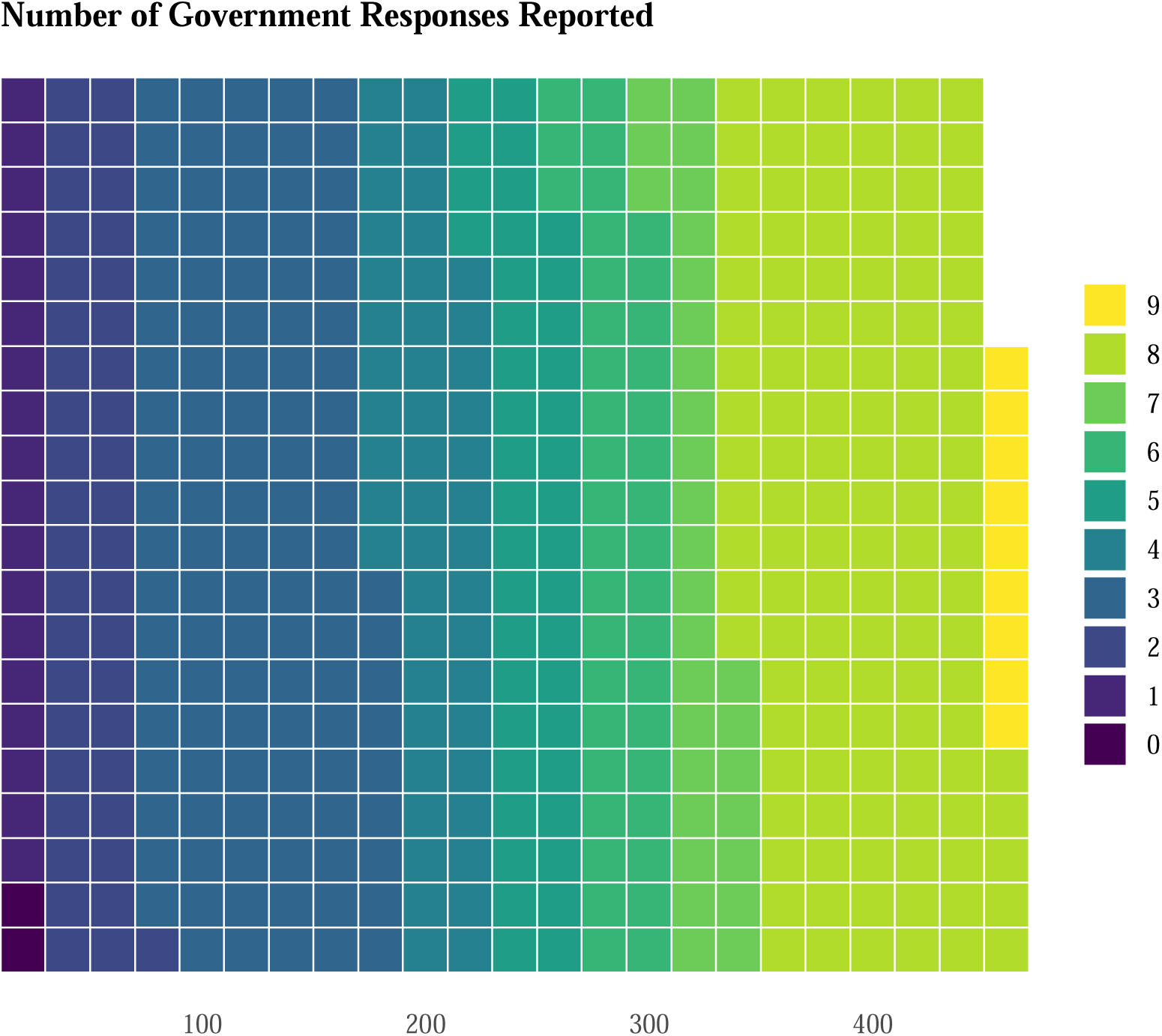
Pattern of Government Response to COVID-19 (n = 454). Counts reflect a summation of all government responses reported by a given household; i.e., advising to avoid gatherings, advising citizens to stay home, travel restrictions, closure of schools, establishing curfews or lockdowns, increasing public awareness, establishing isolation centers, disinfection of public places, closure of non-essential businesses, and other responses.

**Table 1-1:**
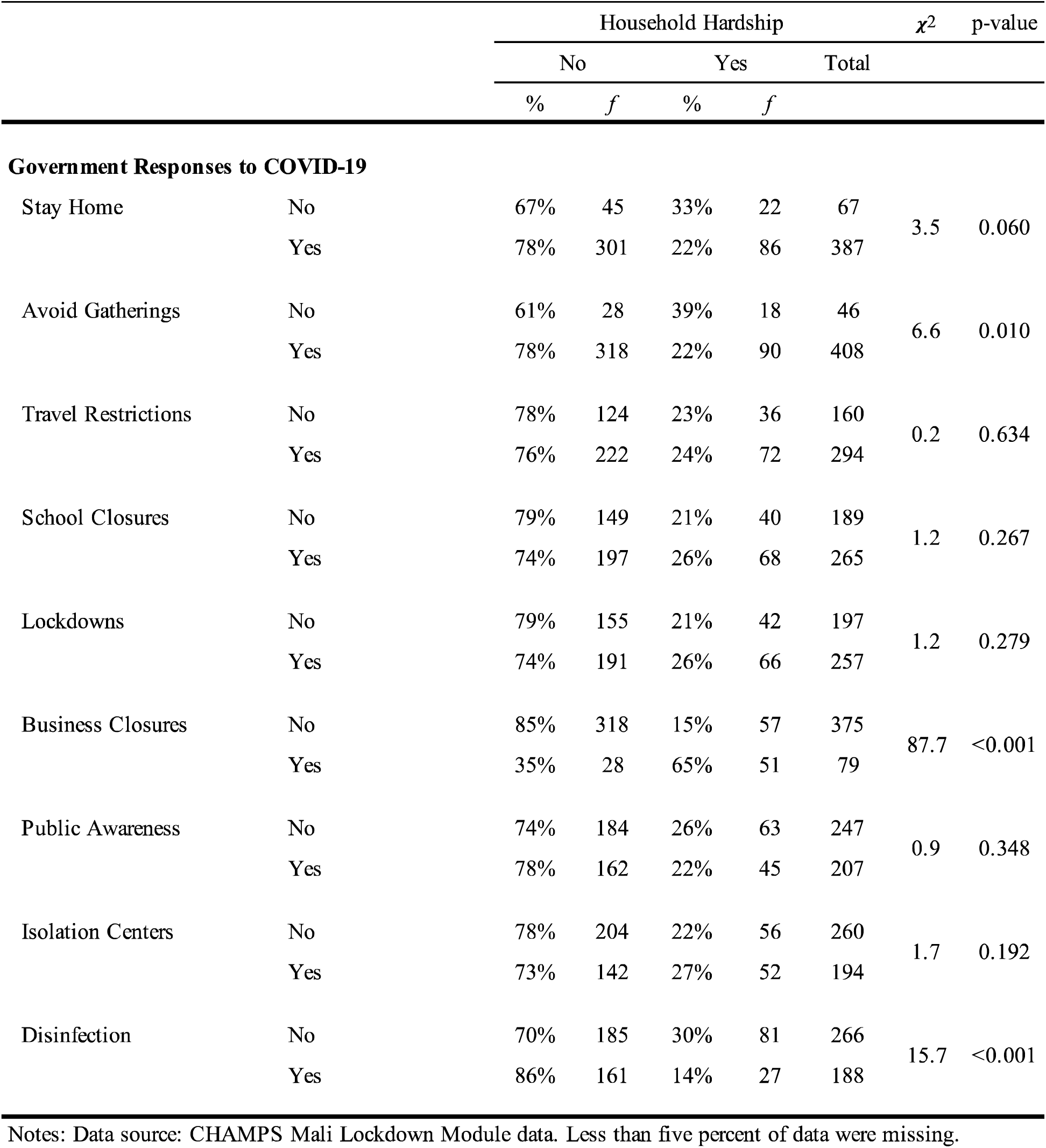
Descriptive Statistics for Government Responses to COVID-19 (n = 454)

After adjusting for other characteristics (see Figure 8), households were more likely to report experiencing at least one hardship during the pandemic if the household head was less than 40 years old (AOR = 3.36, 95% CI [1.23, 9.32]) or between 40 to 49 years old (AOR = 3.06, 95% CI [1.30, 7.35]) compared to 60 years old or older, was male (AOR = 3.91, 95% CI [1.27, 12.50]) compared to female, was an “Other” ethnic group compared to Bambara (AOR = 4.42, 95% CI [1.88, 10.64]) or if their residence was located in Banconi as compared to a Djicoroni (AOR 6.17, 95% CI [2.13, 20.17]). Compared to households with 1 to 4 household members, larger households were more likely to have reported experiencing at least one hardship, but these differences were not statistically significant. Finally, after adjusting for other characteristics, households were more likely to have reported experiencing at least one hardship if the government had closed businesses in the local area (AOR = 18.62, 95% CI [8.62, 42.11]). Conversely, households were less likely to report experiencing at least one hardship during the pandemic if the household head’s occupation was reported as professional (AOR = 0.21, 95% CI [0.06, 0.72]), sales (AOR = 0.40, 95% CI [0.16, 0.97]), or manual labor (AOR = 0.39, 95% CI [0.16, 0.97]). It is noteworthy that in an unadjusted analysis, households were less likely to have reported experiencing at least one hardship if the head of household had either a primary (OR = 0.41, 95% CI [0.23, 0.72]) or secondary/post-secondary (OR = 0.51, 95% CI [0.27, 0.91]) level education compared to no education; however, after adjusting for other variables, this association was no longer statistically significant. Households were also less likely to have reported experiencing at least one hardship if the government had directed citizens in the local area to stay home (AOR = 0.10, 95% CI [0.03, 0.31]) or had taken efforts to disinfect public areas (AOR = 0.23, 95% CI [0.10, 0.51]). As noted above, interaction terms included in statistical models to assess potential effect modification of association based on location of household residence (Banconi vs. Djicoroni) did not yield statistically significant results.

**Figure 8.**
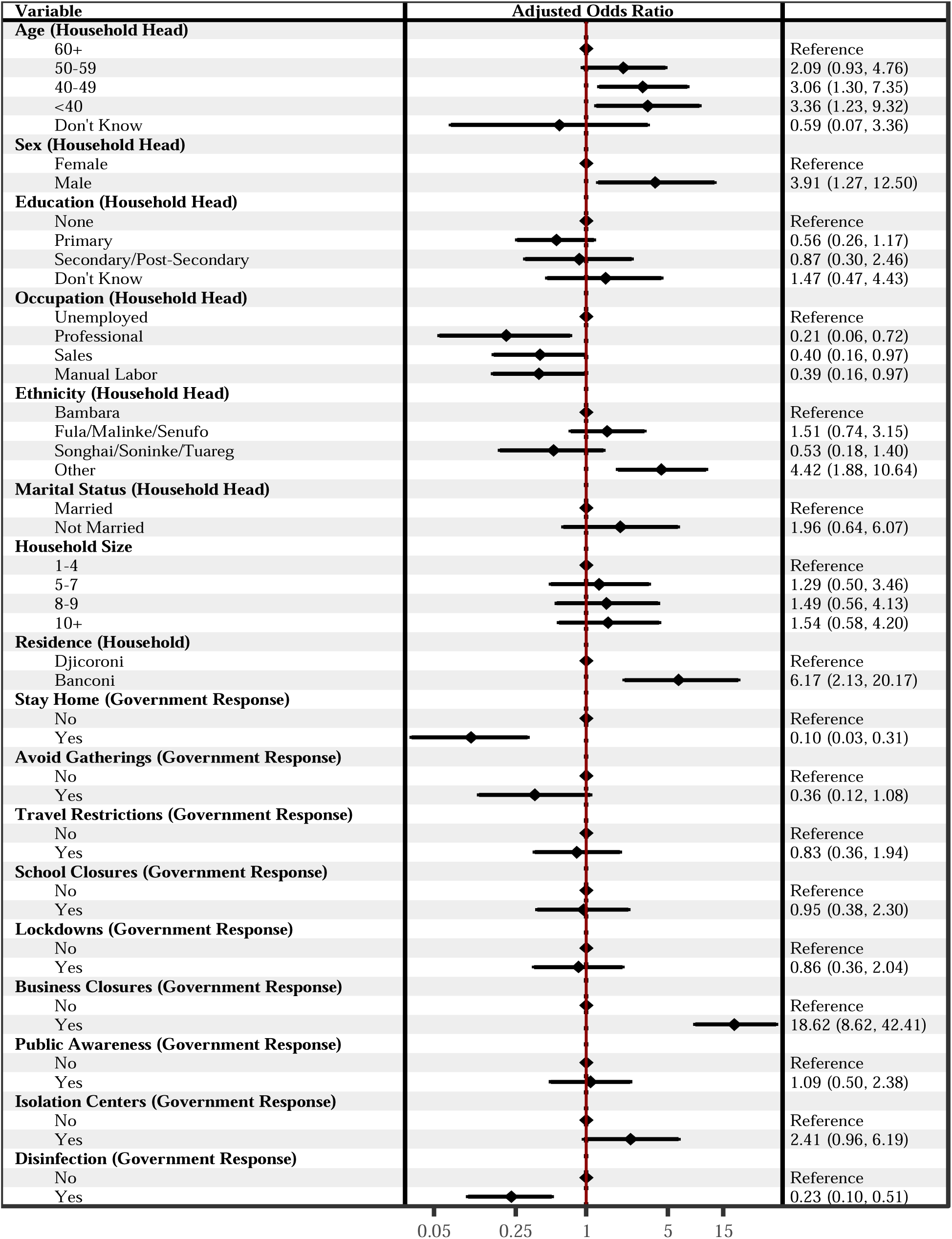
Adjusted Associations between Individual and Household Characteristics with Hardships Reported by Households Living in Bamako, Mali (n=454). Presented as Adjusted Odds Ratios with 95% confidence intervals. Values were missing for less than one percent of all observations.

## DISCUSSION

In this study, we examined the prevalence of households experiencing any of a variety of hardships during the COVID-19 pandemic in Bamako, Mali. We further examined demographic, economic, and social characteristics of individuals and households associated with these hardships.

A little less than 20% of households reported experiencing at least one hardship since the onset of the pandemic, the most commonly reported hardship was an increase in local food prices. This is somewhat lower than anticipated compared to similar studies conducted in other countries. (6, 7, 8) This may reflect the degree to which challenging circumstances are already omnipresent in Mali; e.g., we found that among households residing in the two Bamako areas almost half of household heads had no education and 1 in 5 reported their occupation as *unemployed*, findings that are consistent with WHO findings that adult illiteracy is approximately 65 percent.(36) Individual-level risk factors for experiencing these hardships included head of household characteristics such as younger age, male, and unemployment. At the household level, location of residence was likewise a risk factor. These associations are largely consistent with risk factors reported in studies from other resource-limited countries.(45, 46, 47, 48) Future interventions that seek to mitigate negative consequences associated with pandemic mitigation efforts should anticipate these characteristics as important risk factors and target interventions to aid vulnerable groups.

The empirical findings from this study are specific to the community living within the study site; a limitation common to studies using HDSS data.(49) In addition, the relatively low overall prevalence of reported hardships limited the ability to comprehensively explore associations between sociodemographic characteristics and specific hardships experienced by households. However, the pattern of our findings are generally consistent with findings reported in other studies from sub-Saharan African countries.(6, 7, 8) As an observational study, other potential limitations include unmeasured variable bias (we did not gather information related to food insecurity beyond increases in local food prices such as severity or duration of food insecurity which might underestimate the true impact of hardships; we also did not collect data on potential psychological stress or mental health impacts) or recall bias due to the extended length of time considered in the study (i.e., some respondents may have forgotten hardships that occurred closer to the onset of the pandemic). In preliminary analyses, we considered evaluating additive and Principal Component Analysis (PCA) based indices of these household hardships; however, we concluded that the combination of these variables resulted in a poor fit for an index after evaluating the additive index via Cronbach’s Alpha scores and the results of the PCA analysis. We cannot draw causal inferences given the cross-sectional nature of the study. Results from other studies suggest some hardships associated with the COVID-19 pandemic are ephemeral.(50) Looking forward, scholars could collect and analyze longitudinal data to evaluate hardship events over time to assess the extent to which some hardships may have lingering effects or effects that change over time and explore potential causal relationships. Demographic surveillance systems present an efficient platform for follow-up data collection using the same survey instrument attached to subsequent rounds of regular data collection. Households reported relying almost exclusively on assistance from family and friends as coping mechanism during the pandemic; in future research, scholars could formally explore the role and impact of financial remittances as a specific form of assistance provided across social relationships.

Households in the Bamako HDSS reported experiencing a variety of hardships during the COVID-19 pandemic, with the most prevalent hardships being the increase in local food prices, food insecurity, and job loss. Government closure of businesses was strongly associated with households at risk of experiencing a hardship during the observational period—this points to the need for balancing public health measures with economic considerations during pandemics. Households headed by individuals with lower education and/or unemployed were generally at greater risk of experiencing a hardship. Future policies and practices aiming to mitigate the negative consequences of COVID-19 and future disease outbreaks could anticipate households reflecting these characteristics will disproportionately experiencing socioeconomic hardships.

## Data Availability

All data produced are available online at the CHAMPS Population Surveillance Dataverse.

https://dataverse.unc.edu/dataset.xhtml?persistentId=doi:10.15139/S3/MONHOI

## DECLARATIONS

## Ethics approval and consent to participate

This study was conducted according to the guidelines in the Declaration of Helsinki; all procedures involving research study participants, including digital data collection using tablets that were programmed with the corresponding survey instruments, were approved by the Institutional Ethics Review Board for Health (CIBS) approval reference number No Ref:CIBS-CISM/15/2021. Written informed consent was obtained for participants who were able to read and write. For participants who were unable to read or write, the informed consent statement was read, and oral informed consent from the participant was obtained, documented, and witnessed. These procedures for obtaining written or oral informed consent were approved by the Institutional Ethics Review Board for Health (CIBS), a review board affiliated with the National Bioethics Committee for Health (CNBS).

## Consent for publication

All authors provide consent for publication.

## Availability of data and materials

The COVID-19 lockdown dataset analyzed for this study can be found in the CHAMPS Population Surveillance Dataverse [<u>link</u>].(51, 52) Similar data from studies conducted in Ethiopia, Kenya, and Mozambique are described elsewhere and publicly available.(53, 54, 55, 56, 57)

## Disclaimers

The findings and conclusions in this report are those of the authors and do not necessarily represent the views of the US Centers for Disease Control and Prevention.

## Author contributions

S.S., K.K., M.T., and U.U. established the DSS. J.M., U.U., Z.M., S.S., M.T., K.K., and S.C. provided project administration. J.M., U.U., Z.M., A.N., C.S., and S.C. performed study conceptualization and methodology development. J.M., U.U., and M.O.T. performed data curation. J.M., U.U., Z.M., F.K., M.T., M.O.T., and M.K. carried out data cleaning and validation. J.M. wrote the original draft. All authors reviewed and edited the manuscript. S.S., M.T., K.K., and S.C provided supervision for the study.

## Acknowledgements

We are indebted to the households living within the Bamako HDSS that participated in our study and voluntarily contributed their time and energy in responding to our survey questionnaire in addition to ongoing participation in the regular data collection activities of the HDSS. We are also indebted to the HDSS team members who fielded the survey.

## References

1. Bonaccorsi G, Pierri F, Cinelli M, Flori A, Galeazzi A, Porcelli F, et al. Economic and social consequences of human mobility restrictions under COVID-19. Proceedings of the National Academy of Sciences. 2020;117(27):15530–5.

2. Deb P, Furceri D, Ostry JD, Tawk N. The Economic Effects of COVID-19 Containment Measures. Open Economies Review. 2022;33(1):1–32.

3. Giuntella O, Hyde K, Saccardo S, Sadoff S. Lifestyle and mental health disruptions during COVID-19. Proc Natl Acad Sci U S A. 2021;118(9).

4. Nicola M, Alsafi Z, Sohrabi C, Kerwan A, Al-Jabir A, Iosifidis C, et al. The socio-economic implications of the coronavirus pandemic (COVID-19): A review. Int J Surg. 2020;78:185–93.

5. Verschuur J, Koks EE, Hall JW. Global economic impacts of COVID-19 lockdown measures stand out in high-frequency shipping data. PLoS One. 2021;16(4):e0248818.

6. Muir JA, Dheresa M, Madewell ZJ, Getachew T, Daraje G, Mengesha G, et al. Household hardships and responses to COVID-19 pandemic-related shocks in Eastern Ethiopia. BMC Public Health. 2023;23(1):2086.

7. Nhacolo AQ, Muir JA, Madewell ZJ, Keiri F, Sacoor CN, Jamisse EL, et al. Assessing Sociodemographic Factors Associated with Household Hardships during the COVID-19 Pandemic in Manhiça, Mozambique using Data Collected between April 2021 and February 2022. medRxiv. 2023:2023.12.21.23300355.

8. Muir JA, Dheresa M, Madewell ZJ, Getachew T, Daraje G, Mengesha G, et al. Prevalence of food insecurity amid COVID-19 lockdowns and sociodemographic indicators of household vulnerability in Harar and Kersa, Ethiopia. BMC Nutrition. 2024;10(1):7.

9. Cutter SL, Boruff BJ, Shirley WL. Social vulnerability to environmental hazards. Hazards vulnerability and environmental justice. New York: Routledge; 2012. p. 143–60.

10. Spielman SE, Tuccillo J, Folch DC, Schweikert A, Davies R, Wood N, et al. Evaluating Social Vulnerability Indicators: Criteria and Their Application to the Social Vulnerability Index. Natural Hazards. 2020;100(1):417–36.

11. Muir JA. Societal Shocks as Social Determinants of Health. Columbus: The Ohio State University; 2021.

12. Muir JA, Cope MR, Angeningsih LR, Jackson JE. To Move Home or Move On? Investigating the Impact of Recovery Aid on Migration Status as a Potential Tool for Disaster Risk Reduction in the Aftermath of Volcanic Eruptions in Merapi, Indonesia. International Journal of Disaster Risk Reduction. 2020;46:101478.

13. Muir JA, Cope MR, Angeningsih LR, Jackson JE, Brown RB. Migration and Mental Health in the Aftermath of Disaster: Evidence From MT. Merapi, Indonesia. International journal of environmental research and public health. 2019;16(15):2726.

14. Perry RW. What is a Disaster? Handbook of Disaster Research. New York: Springer; 2007. p. 1–15.

15. Cutter SL, Barnes L, Berry M, Burton C, Evans E, Tate E, et al. A place-based model for understanding community resilience to natural disasters. Global environmental change. 2008;18(4):598–606.

16. Flanagan BE, Gregory EW, Hallisey EJ, Heitgerd JL, Lewis B. A Social Vulnerability Index for Disaster Management. Journal of Homeland Security and Emergency Management. 2011;8(1):1–22.

17. Muir JA. Another mHealth? Examining Motorcycles as a Distance Demolishing Determinant of Health Care Access in South and Southeast Asia. Journal of Transport & Health. 2018;11:153–66.

18. Muir JA. Indicators of Fertility Change in a Developing Nation: Examining the Impact of Motorcycles as a Distance Demolishing Technology on Fertility Change in Rural Indonesia. Provo: Brigham Young University; 2012.

19. Sanders SR, Muir JA, Brown RB. Overcoming Geographic Penalties of Inequality: The Effects of Distance-Demolishing Technologies on Household Well-Being in Vietnam. Asian Journal of Social Science. 2018;46(3):260–80.

20. Muir JA, Sanders SR, Hendricks HZ, Cope MR. Rural Residence, Motorcycle Access, and Contraception Use in South and Southeast AsialZI. Rural Sociology. 2024;89(1):40–62.

21. Goshu D, Ferede T, Diriba G, Ketema M. Economic and Welfare Effects of COVID-19 and Responses in Ethiopia: Initial Insights. Ethiopia: Ethiopian Economic Policy Research Institute; 2020. Contract No.: ISBN 978-99944-54-74-7

22. Bonnet E, Bodson O, Le Marcis F, Faye A, Sambieni NE, Fournet F, et al. The COVID-19 pandemic in francophone West Africa: from the first cases to responses in seven countries. BMC Public Health. 2021;21(1):1490.

23. Adjognon GS, Bloem JR, Sanoh A. The coronavirus pandemic and food security: Evidence from Mali. Food Policy. 2021;101:102050.

24. Bo X. Mali reports first 2 confirmed cases of COVID-19 2020 [cited 2024 February 6]. Available from: http://www.xinhuanet.com/english/2020-03/25/c_138916218.htm.

25. The World Bank. Mali 2024 [cited 2024 February 5]. Available from: https://www.worldbank.org/en/programs/sahel-adaptive-social-protection-program-trust-fund/country-work/mali.

26. United Nations Development Programme. Mali: Human Development Index 2024 [cited 2024 February 5]. Available from: https://hdr.undp.org/data-center/specific-country-data#/countries/MLI.

27. Kawuki J, Chan PS, Fang Y, Chen S, Mo PKH, Wang Z. Knowledge and Practice of Personal Protective Measures Against COVID-19 in Africa: Systematic Review. JMIR Public Health Surveill. 2023;9:e44051.

28. Matovu JK, Mulyowa A, Akorimo R, Kirumira D. Knowledge, risk-perception, and uptake of COVID-19 prevention measures in sub-Saharan Africa: a scoping review. Afr Health Sci. 2022;22(3):542–60.

29. Mudenda S, Mukosha M, Godman B, Fadare JO, Ogunleye OO, Meyer JC, et al. Knowledge, Attitudes, and Acceptance of COVID-19 Vaccines among Secondary School Pupils in Zambia: Implications for Future Educational and Sensitisation Programmes. Vaccines. 2022;10(12):2141.

30. Naidoo D, Meyer-Weitz A, Govender K. Factors Influencing the Intention and Uptake of COVID-19 Vaccines on the African Continent: A Scoping Review. Vaccines (Basel). 2023;11(4).

31. Song S, Madewell ZJ, Liu M, Longini IM, Yang Y. Effectiveness of SARS-CoV-2 vaccines against Omicron infection and severe events: a systematic review and meta-analysis of test-negative design studies. Front Public Health. 2023;11:1195908.

32. Shapiro J, Dean NE, Madewell ZJ, Yang Y, Halloran ME, Longini I. Efficacy estimates for various COVID-19 vaccines: what we know from the literature and reports. Medrxiv. 2021:2021.05. 20.21257461.

33. Dheresa M, Madewell ZJ, Muir JA, Getachew T, Daraje G, Mengesha G, et al. Factors Influencing Knowledge of COVID-19 Prevention in Eastern Ethiopia. SAGE Open. 2024;14(3):21582440241273871.

34. Nhacolo A, Madewell ZJ, Muir JA, Sacoor C, Xerinda E, Matsena T, et al. Knowledge of COVID-19 symptoms, transmission, and prevention: Evidence from health and demographic surveillance in Southern Mozambique. PLOS Global Public Health. 2023;3(11):e0002532.

35. Cunningham SA, Shaikh NI, Nhacolo A, Raghunathan PL, Kotloff K, Naser AM, et al. Health and Demographic Surveillance Systems within the Child Health and Mortality Prevention Surveillance Network. Clinical Infectious Diseases. 2019;69(Supplement_4):S274–S9.

36. Organization WH. The global action plan for healthy lives and well-being for all: strengthening collaboration among multilateral organizations to accelerate country progress on the health-related sustainable development goals [brochure]. World Health Organization; 2019.

37. Ruel E, Wagner III WE, Gillespie BJ. The practice of survey research: Theory and applications. New York: Sage Publications; 2015.

38. World Bank Group. High Frequency Mobile Phone Surveys of Households to Assess the Impacts of COVID-19 (Vol. 4) : Questionnaire Template (English) Washington, D.C.2020 [cited 2024 February 6]. Available from: http://documents.worldbank.org/curated/en/567571588697439581/Questionnaire-Template.

39. Harris PA, Taylor R, Minor BL, Elliott V, Fernandez M, O’Neal L, et al. The RedCAP Consortium: Building an International Community of Software Platform Partners. Journal of Biomedical Informatics. 2019;95:103208.

40. Cunningham S, Muir J, Nichols A, Edlund J. Data Cleaning. The Cambridge Handbook of Research Methods and Statistics for the Social and Behavioral Sciences. 2023;1:443–67.

41. Van den Broeck J, Argeseanu Cunningham S, Eeckels R, Herbst K. Data cleaning: detecting, diagnosing, and editing data abnormalities. PLoS medicine. 2005;2(10):e267.

42. R Core Team. R: A Language and Environment for Statistical Computing. Vienna, Austria: R Foundation for Statistical Computing; 2022.

43. Yeh J, D’Amico F. Forest plots: data summaries at a glance. Journal of Family Practice. 2004;53(12):1007–8.

44. Lewis SC, Keerie C, Assi V. Forest Plot. Wiley StatsRef: Statistics Reference Online2014. p. 1–6.

45. Onyango EO, Crush J, Owuor S. Preparing for COVID-19: Household Food Insecurity and Vulnerability to Shocks in Nairobi, Kenya. PLoS ONE. 2021;16(11):e0259139.

46. Fikire AH, Zegeye MB. Determinants of Rural Household Food Security Status in North Shewa Zone, Amhara Region, Ethiopia. The Scientific World Journal. 2022;2022:1–8.

47. Manfrinato CV, Marino A, Condé VF, Maria do Carmo PF, Stedefeldt E, Tomita LY. High prevalence of food insecurity, the adverse impact of COVID-19 in Brazilian favela. Public health nutrition. 2021;24(6):1210–5.

48. Banna M, Al H, Sayeed A, Kundu S, Kagstrom A, Sultana M, et al. Factors associated with household food insecurity and dietary diversity among day laborers amid the COVID-19 pandemic in Bangladesh. BMC nutrition. 2022;8(1):1–11.

49. Clark S, Wakefield J, McCormick T, Ross M. Hyak Mortality Monitoring System: Innovative Sampling and Estimation Methods–Proof of Concept by Simulation. Global Health, Epidemiology and Genomics. 2018;3:1–14.

50. Hirvonen K, de Brauw A, Abate GT. Food Consumption and Food Security During the COVID-19 Pandemic in Addis Ababa. American Journal of Agricultural Economics. 2021;103(3):772–89.

51. Omwuchekkwa U, Traore M, Muir J, Madewell Z, Keiri F, Cunningham S, et al. COVID-19 Impact Data for the CHAMPS HDSS Network: Data from Bamako, Mali. V1 ed: UNC Dataverse; 2023.

52. Muir JA, Onwuchekwa UU, Madewell ZJ, Traore MO, Kourouma M, Keiri F, et al. Coronavirus Awareness, Prevention, and Household Hardship Survey Data for the CHAMPS HDSS Network: Data Collected between August and September of 2022 from the Bamako HDSS, Mali. Data in Brief. 2024:110651.

53. Dheresa M, Muir JA, Madewell ZJ, Getachew T, Daraje G, Mengesha G, et al. COVID-19 Impact Data for the CHAMPS HDSS Network: Data from Harar and Kersa, Ethiopia. V1 ed: UNC Dataverse; 2023.

54. Muir JA, Dheresa M, Madewell ZJ, Getachew T, Daraje G, Mengesha G, et al. COVID-19 impact data for the CHAMPS HDSS network: Data from Harar and Kersa, Ethiopia. Data in Brief. 2023;50:109508.

55. Nhacolo A, Sacoor C, Xerinda E, Matsena T, Muir JA, Madewell ZJ, et al. COVID-19 Impact Data for the CHAMPS HDSS Network: Data from Manhiça, Mozambique. V1 ed: UNC Dataverse; 2023.

56. Muir JA, Matsena T, Madewell ZJ, Keiri F, Sacoor CN, Jamisse EL, et al. Coronavirus Awareness and Household Hardship Survey Data for the CHAMPS HDSS Network: Data Collected between April 2021 and February 2022 in the Manhiça HDSS, Mozambique. Data in Brief. 2024:110654.

57. Obor D, Muir JA, Munga S, Barr BT, Khaggayi C, Awiti L, et al. COVID-19 Impact Data for the CHAMPS HDSS Network: Data from Karemo and Manyatta, Kenya. V1 ed: UNC Dataverse; 2023.

